# Resistance and prevalence implications of doxycycline post-exposure prophylaxis for gonorrhea prevention in men who have sex with men: a modeling study

**DOI:** 10.1101/2023.04.24.23289033

**Authors:** Emily Reichert, Yonatan H. Grad

## Abstract

**Background:** Doxycycline post-exposure prophylaxis (DoxyPEP) has demonstrated efficacy for prevention of bacterial sexually transmitted infections. To inform policy decisions on the use of DoxyPEP for gonorrhea prevention, we used a mathematical model to investigate its impact on resistance dynamics and the burden of infection in men who have sex with men (MSM).

**Methods and Findings:** Using a deterministic compartmental model of gonorrhea transmission in an MSM population, we introduced DoxyPEP at various uptake levels (10-75%) and compared 20-year prevalence and resistance dynamics relative to those at baseline (i.e., no DoxyPEP introduction). Uptake of DoxyPEP resulted in initial drops in the prevalence and incidence of gonorrhea infection, but also accelerated the spread of doxycycline resistance, with increasing DoxyPEP use driving steeper initial declines followed by faster spread of resistance. This resulted in the total loss of DoxyPEP’s clinical efficacy within 1-2 decades in almost all scenarios explored. The magnitude by which DoxyPEP initially reduced the prevalence of infection was constrained by the extent of pre-existing doxycycline resistant strains in the population. *De novo* emergence of doxycycline resistance did not influence these dynamics. Additionally, the implementation of DoxyPEP had minimal impact on extending the clinically useful lifespan of ceftriaxone monotherapy.

**Conclusions:** Model findings suggest DoxyPEP can be an effective but short-term solution for reducing the burden of gonorrhea infection, as its selection for doxycycline-resistant strains results in loss of its prophylaxis benefit. Increasing levels of DoxyPEP uptake and higher starting prevalence of doxycycline resistance resulted in faster loss of its efficacy and had little change on extending the clinical lifespan of ceftriaxone for treatment of *N. gonorrhoeae* infections.

## Introduction

Doxycycline post-exposure prophylaxis (known as DoxyPEP), a 200mg dose of the broad-spectrum antibiotic doxycycline after condomless sexual contact, shows evidence in some clinical trials of reducing the incidence of bacterial sexually transmitted infections (STIs) in high-risk populations. While risk reductions have been consistent for chlamydia and syphilis across geographies, results for gonorrhea have been mixed. Findings from a substudy of the ANRS IPERGAY trial, enrolling 232 men who have sex with men (MSM) in France, showed no significant difference in the time until first *Neisseria gonorrhoeae* (also known as gonococcus) infection among DoxyPEP users vs. non-users (hazard ratio [HR] = 0.83; p = 0.52) (1). In contrast, the DoxyPEP trial conducted in the United States (U.S.) of 501 high-risk MSM and transgender women (TGW) in Seattle and San Francisco demonstrated a significant reduction in the quarterly cumulative incidence of gonococcal infection in the DoxyPEP arm relative to the control arm for both those living with HIV (risk ratio [RR; 95% CI] = 0.43 [0.26-0.71]) and using pre-exposure prophylaxis (PrEP) for HIV (0.45 [0.32-0.65]) (2). Differences in the efficacy of DoxyPEP in preventing gonococcal infection by geography may be due to underlying variation in the population-level prevalence of resistance to the broader tetracycline class; 56% of gonococcal isolates were resistant to tetracyclines in France in 2012, and in Kenya, the site of an ongoing DoxyPEP trial, 96% of isolates collected from women in 2008-2012 were resistant (3,4).

Despite the lack of national guidance on its use in the U.S., interest in DoxyPEP is high, particularly among the MSM population. One racially-ethnically and geographically diverse U.S. survey found 84% of users of a gay social-networking application interested in DoxyPEP (5). The San Francisco Department of Public Health is among the first globally to formally recommend DoxyPEP for high-risk cis men and trans women, whereas others such as the United Kingdom Health Security Agency (UKHSA) cite antimicrobial resistance concerns as reason to not endorse DoxyPEP (6). Without a formal endorsement, the U.S. Centers for Disease Control and Prevention (CDC) acknowledges that some clinicians and individuals utilize DoxyPEP off-label, and they include considerations for DoxyPEP within STI treatment guidelines as they continue “to evaluate data to inform clinical guidance” (7). To inform this policy decision – whether to recommend DoxyPEP use despite concerns about selection for *N. gonorrhoeae* resistance – we explored the effect of DoxyPEP implementation on gonococcal infection and resistance dynamics in a large population through a mathematical model of gonorrhea transmission among MSM.

## Methods

### Model Overview

We adapted a deterministic compartmental model characterizing gonorrhea transmission in a U.S. MSM population (8). We added an exposure compartment to study the dynamics of administering doxycycline post-exposure prophylaxis (DoxyPEP) to a proportion of exposed individuals (ξ_B_), transforming the model into a susceptible-exposed-infectious-susceptible (SEIS) model (Supplementary Figure 1). Individuals spent on average 1 day in the exposure (or latent) compartment, after which they either progressed to infection or transitioned back to susceptibility following successful doxycycline prophylaxis. The rate of removal from the exposed compartment (γ = 1/(1 day)) was in line with the recommended 72 hour window for DoxyPEP following the exposure event (9).

In brief, the model characterized an MSM population (N = 10^6^) stratified into 3 sexual activity groups characterized by annual rates of partner change (θ), with individuals of different sexual activity groups interacting with mixing parameter ε. Individuals aged into and out of the sexually active population at rate ρ, contributing for 20 years on average. Infected individuals could recover spontaneously or through antibiotic treatment with ceftriaxone monotherapy. Infections were stratified by symptomatic (denoted Y) versus asymptomatic (Z) status and by resistance profile, where each infection could be resistant to ceftriaxone (denoted A), doxycycline (B), neither, or both.

### Model Parameterization

We parameterized the model using the literature and the model fitting procedure, which used maximum likelihood estimation (MLE) to fit the model to a mean 3.0% (variance 1.47 × 10^−5^) baseline equilibrium prevalence of gonorrhea infection based on recent surveillance estimates in MSM (10,11). The calibration SEIS model with no DoxyPEP implementation was run over two years, and parameter MLEs were calculated using R package bbmle (12). Model parameters are listed in Supplementary Table 1.

Isolates from the 2018 Gonococcal Isolate Surveillance Project (GISP) were used to inform the proportion of doxycycline resistant infections. As tetracycline, but not doxycycline, resistance is reported, the prevalence of tetracycline resistance in MSM (0.268, or 26.8%) was used as a proxy measure (13). To account for uncertainty and to allow generalizability of results to other geographic settings, we vary this initial prevalence of doxycycline resistance in sensitivity analysis (0.20-0.80). Estimates from 2020-2021 GISP data in MSM were used to inform the starting prevalence of ceftriaxone resistance (0.01%) (14).

Parameters of ceftriaxone, including the probability of *de novo* resistance (ω_A_) and the fitness cost associated with resistance (1-f_A_), were inferred from the literature (15,16). Doxycycline resistant strains were assumed to have the same small fitness cost (2%) as ceftriaxone resistant strains in our primary analysis; sensitivity analyses explored outcomes over a wider range of potential fitness costs (1-f_B_: 0-20%). In the primary analysis, we assumed no emergence of doxycycline resistance on DoxyPEP treatment (i.e., *de novo* resistance emergence was 0%), but this also varied in sensitivity analyses (ω_B_: 0-10^−4^). While we refer to our two modeled drugs as ceftriaxone and doxycycline, parameters are only informed estimates of these drugs’ probabilities of *de novo* resistance and resistance-associated fitness costs, which are inherently difficult properties to observe and measure in vivo. Ceftriaxone monotherapy was assumed to be the treatment given to 100% of those who progressed to gonococcal infection; those with symptomatic ceftriaxone resistant infections (for which this therapy is ineffective) could seek retreatment at rate T_sr_ with probability π_s_. For these cases, we assigned retreatment with a last-resort antibiotic external to our model for which we assumed complete efficacy and did not monitor resistance trends.

DoxyPEP therapy was assumed to be unsuccessful in preventing infection caused by exposure to a doxycycline resistant strain. Data from the DoxyPEP trial showed a 55% reduced quarterly cumulative incidence of gonococcal infection for MSM and TGW DoxyPEP users (RR = 0.45) compared to non-users (2). Therefore, assuming that 26.8% of those receiving DoxyPEP had risk ratio [RR] = 1 due to pre-existing doxycycline resistance, we used a weighted average to calculate the RR of those exposed to doxycycline susceptible strains that would allow us to observe an overall RR = 0.45. This implied the quarterly relative risk of infection for the remaining 73.2% of the population with exposure to susceptible bacteria was 0.25 for DoxyPEP users relative to non-users, consistent with the range of efficacies in preventing chlamydia and syphilis reported for the DoxyPEP trial (2). For the primary analysis, we made the simplifying and optimistic assumption that this corresponded to a 75% reduction in the risk of infection *per exposure event* (i.e., the proportion of DoxyPEP treatments that failed to prevent gonococcal infection for reasons not related to resistance was κ = 0.25). This holds if individuals in the DoxyPEP study were exposed to *N. gonorrhoeae* once on average during a three month period. To test the impact of this assumption, we explored a range of risk ratios per exposure event (κ: 0-0.8) in sensitivity analyses.

### Model Implementation

We ran the model over 20 years using R package deSolve (17) to observe the projected dynamics of ceftriaxone and doxycycline resistance, as well as the burden of gonococcal infection, following DoxyPEP implementation at t = 0. A range of potential uptake, or utilization, levels were explored (0, 10, 25, 50, and 75%) that characterized the proportion of exposures to gonococcal infection administered DoxyPEP as intended. We evaluated multiple primary outcomes over time following DoxyPEP introduction, including: the prevalence, incidence rate, and cumulative number of gonococcal infections; the cumulative number of ceftriaxone treatments administered; the time until 5% resistance prevalence for ceftriaxone, 5% resistance prevalence for dual resistance, and 87% resistance prevalence for doxycycline. We chose a threshold of 5% resistance prevalence for ceftriaxone as this represents the World Health Organization’s threshold for revisiting treatment guidelines (18). For doxycycline, since resistance levels in the U.S. are well above this threshold at baseline (14), we arrived at the threshold of 87% prevalence by calculating the threshold for which there was a less than 10% reduction in risk of infection with DoxyPEP use (i.e., the burden of infection from *N. gonorrhoeae* resistant to doxycycline resulted in an estimated gonorrhea RR of > 0.90 [assuming κ = 0.25] for DoxyPEP users vs. non-users).

In addition to the sensitivity analyses exploring parameter uncertainty (in κ, f_B_, ω_B_, and initial doxycycline resistance prevalence), we conducted an additional analysis that restricted DoxyPEP utilization to the high sexual activity group, in line with a policy that recommends DoxyPEP only for high-risk individuals.

All of the code needed to run the model as well as analyze and visualize results is available at https://github.com/emreichert13/doxypep.

## Results

Under baseline parameterization, the transmission model projected that policy recommendations leading to DoxyPEP uptake at or above 10% would result in a substantial reduction in the prevalence of gonococcal infection over the initial implementation period (Figure 1). Within the first 5 years after DoxyPEP was introduced (at t=0), the prevalence of infection was reduced by up to 62%, with the magnitude of the reduction increasing with the percentage of DoxyPEP uptake. Incidence rates for gonorrhea, measuring the rapidity at which individuals acquire infection, displayed similar trends following DoxyPEP introduction (Supplementary Figure 2). The percent reduction in the cumulative number of infections after 5 years ranged from 19.5% (10% uptake) to 49.7% (75% DoxyPEP uptake), relative to the ceftriaxone monotherapy *status quo* (Table 1). As time since DoxyPEP introduction progressed, and doxycycline resistance became increasingly widespread, its benefit in reducing the burden of infection tapered. After 20 years, differences in the gonococcal infection prevalence across non-zero DoxyPEP uptake levels (≥ 10%) had largely disappeared. The percent reduction in cumulative infections ranged from 13.5% (10% uptake) to 14.6% (75% DoxyPEP uptake) compared with no DoxyPEP use (Table 1). Differences in which resistant strain grew to dominate circulating infections if any level of DoxyPEP (≥ 10%) was implemented, introducing a selective pressure for doxycycline resistance, explain the persistent difference in the gonococcal infection prevalence for the 0% DoxyPEP uptake scenario relative to others even beyond widespread DoxyPEP failure (Figure 1, Supplementary Figure 3).

**Figure 1.**
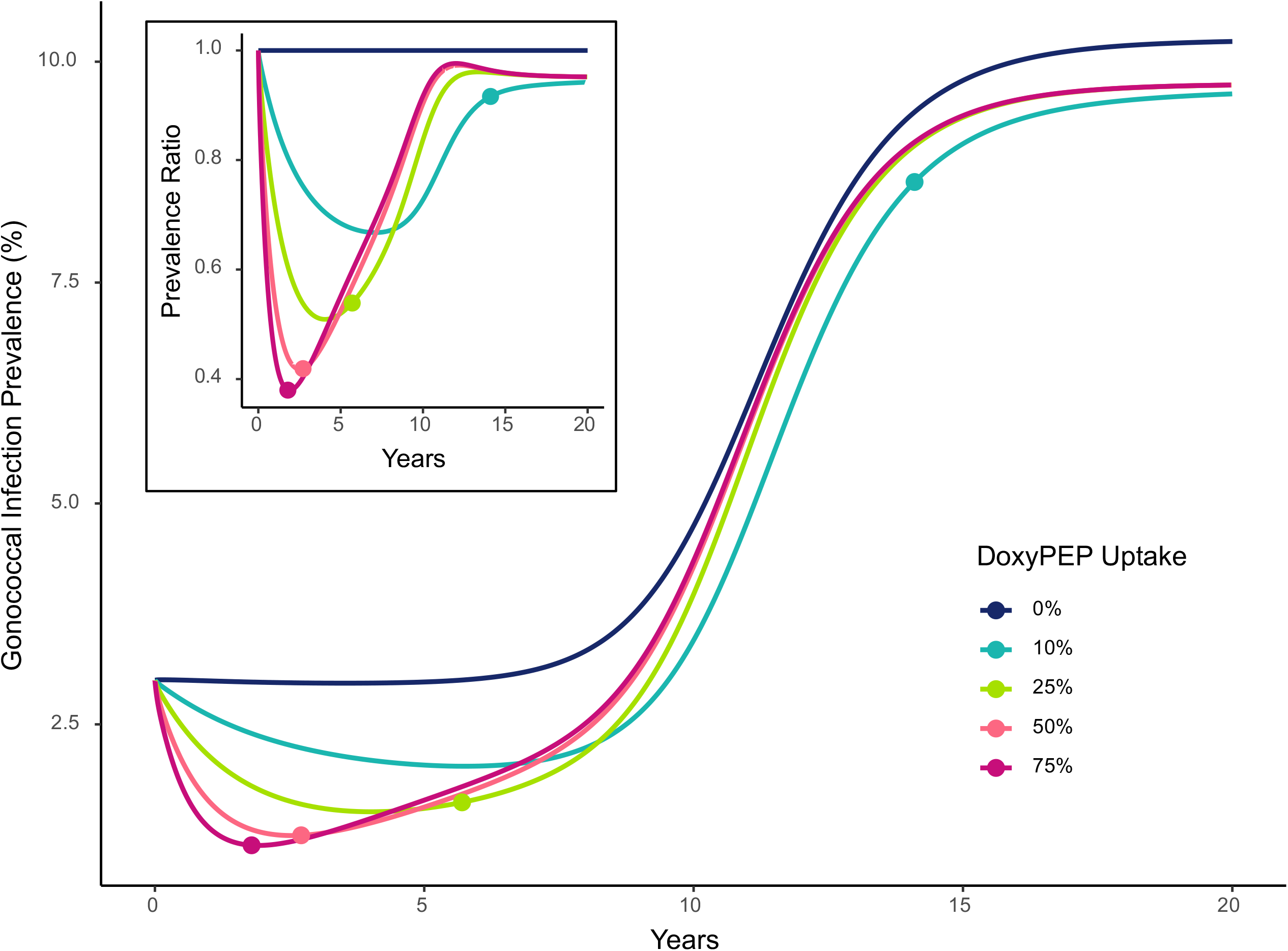
Prevalence of gonococcal infection over time for varying DoxyPEP uptake levels (%). Prevalence at each timepoint is calculated as the total number of infections over the total population size (N = 10^6^). The upper left inset visualizes the same data as a prevalence ratio (PR), where results are normalized (or divided by) the prevalence under the no DoxyPEP introduction scenario (0% uptake). Lines are colored by DoxyPEP uptake, defined as the proportion of exposure events that are treated with doxycycline post-exposure prophylaxis within the population. The points along these lines represent the time at which doxycycline resistance reached 87% prevalence, the threshold for which there was a less than 10% reduction in risk of infection with DoxyPEP use. Abbreviations: DoxyPEP = doxycycline post-exposure prophylaxis.

**Table 1.** Clinically relevant endpoints by level of DoxyPEP uptake (%). Results are calculated using baseline model parameters listed in Supplementary Table 1.

| DoxyPEP Uptake | Time to Ceftriaxone Resistance at 5% (years) | Time to Dual Resistance at 5% (years) | Time to Doxycycline Resistance at 87% (years) | Reduction in Cumulative Ceftriaxone Treatments at 5 years (%), relative to no DoxyPEP uptake | Reduction in Cumulative Ceftriaxone Treatments at 20 years (%), relative to no DoxyPEP uptake | Reduction in Cumulative Infections at 5 years (%), relative to no DoxyPEP uptake | Reduction in Cumulative Infections at 20 years (%), relative to no DoxyPEP uptake |
| --- | --- | --- | --- | --- | --- | --- | --- |
| 0% | 6.6 | 9.1 | NA | NA | NA | NA | NA |
| 10% | 6.7 | 7.2 | 14.1 | 23.0% | 16.0% | 19.5% | 13.5% |
| 25% | 6.5 | 6.5 | 5.7 | 41.5% | 17.4% | 36.5% | 14.4% |
| 50% | 6.3 | 6.3 | 2.7 | 51.7% | 17.6% | 46.6% | 14.5% |
| 75% | 6.3 | 6.3 | 1.8 | 54.7% | 17.6% | 49.7% | 14.6% |

Across DoxyPEP utilization levels we evaluated (10-75%), the prevalence of doxycycline resistance consistently rose to the threshold (87%) at which there was a less than 10% reduction in risk of infection with DoxyPEP use per exposure event within the 20-year period. As DoxyPEP use increased, the time until widespread doxycycline resistance and until strains dually resistant to doxycycline and ceftriaxone achieved 5% prevalence both decreased (Figure 1, Table 1). The introduction of DoxyPEP within the population and its subsequent level of use did not impact the time until ceftriaxone resistance met 5% prevalence (Table 1). High levels of DoxyPEP use (50-75%) reduced the number of ceftriaxone treatments administered over the first five years by > 50% compared to baseline, but after 20 years this reduction had narrowed to 17.6% (Table 1).

We then restricted use of DoxyPEP to those in the highest sexual activity group, comprising a fixed 10% of the population, to assess infection and resistance dynamics following a DoxyPEP rollout strictly targeting those at high risk for STIs. We considered the same range of use (0-75%), but as these only characterize the high-risk group with use fixed at 0% for the remaining 90% of the population, absolute use of DoxyPEP was reduced. Trends in the 20-year prevalence of infection were highly similar to those when the entire population was eligible for DoxyPEP (Supplementary Figure 4). Prevalence of infection was reduced by up to 58% (75% uptake), with the magnitude of the relative reduction in prevalence again increasing with a greater percentage of DoxyPEP uptake. The time to doxycycline resistance reaching the 87% resistance prevalence threshold (at which the expected risk reduction for those on DoxyPEP is <10%) was slightly extended, and at 5 years the percent reduction in ceftriaxone treatments was slightly attenuated (Supplementary Table 2). Other outcomes differed by less than a 20 percentage change from the primary analysis results.

To evaluate the impact of DoxyPEP’s efficacy in preventing infection per exposure event to doxycycline-susceptible strains (captured by risk ratio parameter κ), we varied this efficacy from 20-100% and again assessed the 20-year trends in prevalence of infection under a range of DoxyPEP uptake scenarios (Supplementary Figure 5). As the risk ratio κ increased (i.e., efficacy of DoxyPEP decreased), the benefits of DoxyPEP in reducing the prevalence of infection were attenuated, but the time until doxycycline resistance reached 87% prevalence within each uptake level was extended.

We next evaluated the robustness of results to alternative parameterizations of the fitness cost associated with doxycycline resistance (1-f_B_: 0-0.20) and the probability of doxycycline resistance emerging and establishing resistant infection (ω_B_: 0-10^−4^) in a bivariate sensitivity analysis (Supplementary Figures 6-7). The effectiveness of DoxyPEP in reducing the burden of gonococcal infection and the time until DoxyPEP led to widespread doxycycline resistance were each insensitive to the *de novo* resistance parameter ω_B_ over our explored parameter range. In contrast, varying the relative fitness of doxycycline-resistant strains compared to doxycycline-susceptibles (f_B_) led to qualitatively different gonococcal infection and resistance dynamics. Increasing the fitness cost associated with doxycycline resistance extended the time until doxycycline resistance reached the 87% threshold within each DoxyPEP uptake level. The combination of i) a high DoxyPEP use and ii) a substantial fitness cost associated with doxycycline resistance led to maintenance of a very low burden of gonococcal infection over the 20-year window relative to no DoxyPEP introduction. For example, assuming a 20% fitness cost associated with doxycycline resistance and a DoxyPEP uptake level of ≥ 25%, the 20-year cumulative gonococcal infection count was reduced by ≥ 77.0%; with ≥ 50% DoxyPEP uptake, cumulative infections were reduced by ≥ 91.2% relative to the projected infections over this 20-year period with 0% DoxyPEP use. The underlying resistance dynamics for these model parameterizations show that, while dual resistant strains still rise to comprise ≥ 99% of infections within 20 years if DoxyPEP uptake is ≥ 50%, the drastically reduced fitness of dual resistant strains maintains the low burden of gonorrhea infection even after DoxyPEP loses its clinical utility, or ability to prevent gonococcal infection post-exposure.

Lastly, to evaluate the impact of DoxyPEP in settings with higher baseline prevalence of doxycycline resistance, we reran the model with initial resistance estimates ranging from 0.20-0.80 (Figure 2). With higher resistance to doxycycline at baseline, both the time until widespread doxycycline resistance (>87%) and the clinical benefit of DoxyPEP decreased.

**Figure 2.**
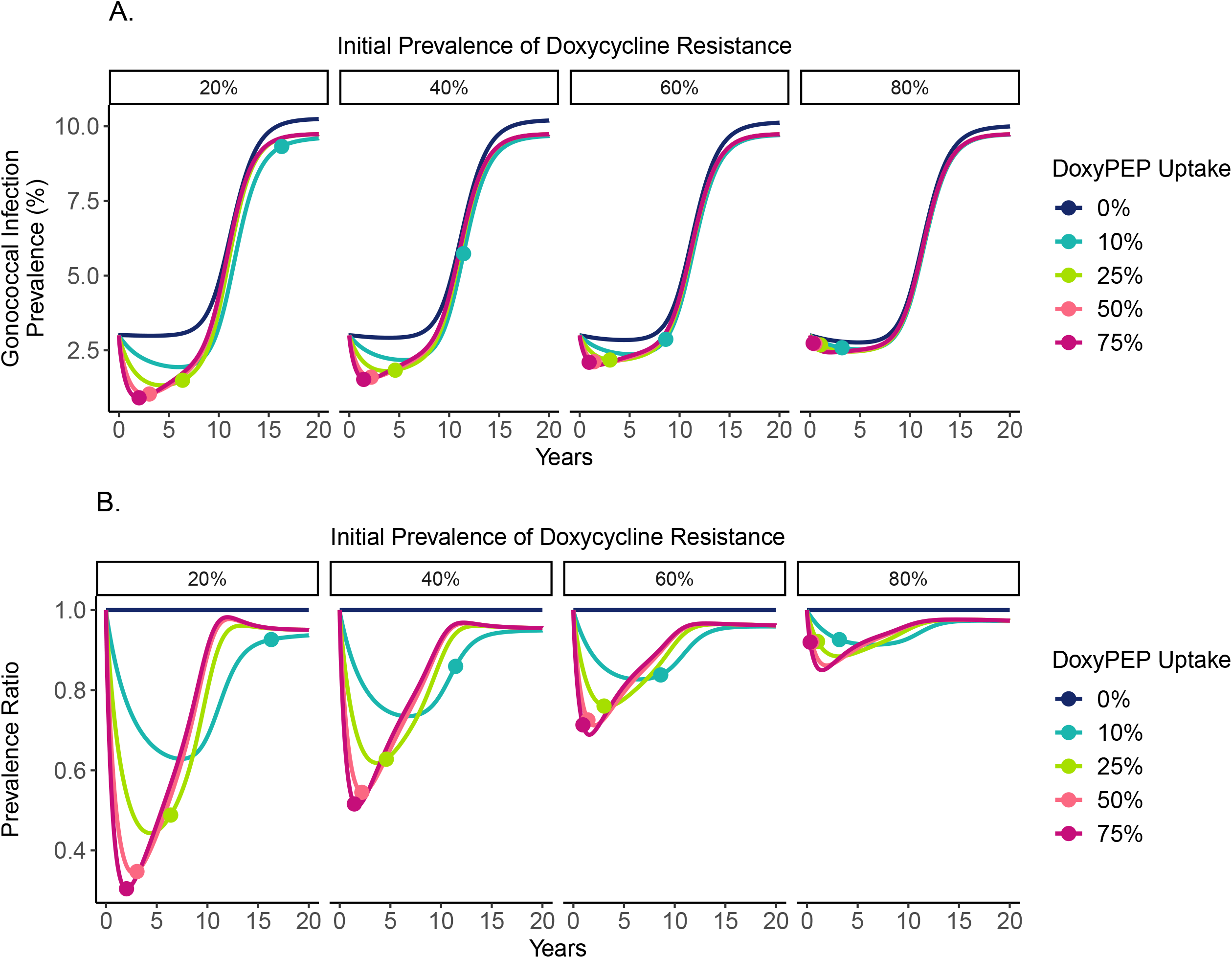
Prevalence of gonococcal infection over time for varying DoxyPEP uptake levels (%), by the prevalence of doxycycline resistance at time 0. **A)** Absolute prevalence estimates over time, calculated as the total number of gonococcal infections over the total population size (N = 10^6^) under each DoxyPEP utilization scenario. **B)** Prevalence ratio (PR) estimates over time, where results are normalized (or divided by) the prevalence under the no DoxyPEP introduction scenario (0% uptake). Vertical facets (columns) show the prevalence of doxycycline resistance among gonococcal infections at the start of the model, ranging from 20-80%. Lines are colored by DoxyPEP uptake, defined as the proportion of exposure events that are treated with doxycycline post-exposure prophylaxis within the population. The points along these lines represent the time at which doxycycline resistance has reached an 87% prevalence among gonococcal infections under that uptake level. Abbreviations: DoxyPEP = doxycycline post-exposure prophylaxis.

## Discussion

Our analysis demonstrated that, under most model parameterizations explored, DoxyPEP implementation was a highly effective but short-term intervention for reducing the burden of gonorrhea infection. As DoxyPEP use increased, so too did its effectiveness in reducing gonorrhea prevalence and incidence, initially reducing prevalence by > 50% in high-uptake scenarios. However, increasing DoxyPEP use also led to accelerated loss of its clinical utility, with the direct prophylactic benefit of DoxyPEP almost always lost within 1-2 decades due to high prevalence of doxycycline resistance. This was not due to the evolution of doxycycline resistance upon DoxyPEP treatment, but instead due to DoxyPEP’s population-level effect of preventing infections by doxycycline-susceptible strains and preferentially selecting for doxycycline-resistant ones. The results demonstrate the classic tension that comes with using antimicrobials between immediate clinical benefit for an individual and potential future cost for a population. More DoxyPEP use led to greater reductions in the prevalence of gonococcal infection but at the cost of accelerated loss of DoxyPEP’s efficacy due to increasing doxycycline resistance.

Under baseline model parameterization, introducing DoxyPEP at any non-zero uptake level leads to 5-year reductions in the cumulative number of ceftriaxone treatments ranging from 23.0-54.7% relative to baseline (Table 1). These relative reductions level off at around 18% across uptake levels by year 20. However, the introduction of DoxyPEP into the model population did not “buy more time” in terms of the prevalence of ceftriaxone resistance. Across DoxyPEP implementation scenarios (entire population vs. high-risk only) and uptake levels (0-75%), the time until 5% ceftriaxone resistance prevalence stayed relatively constant (Table 1, Supplementary Table 2). Of note, this measure only reflects the proportion of infections and not the absolute number that are ceftriaxone-resistant.

Restricting DoxyPEP utilization to only the sexual activity group with the highest partner turnover rate (which comprises 10% of our model population) only slightly reduced DoxyPEP’s benefit in reducing gonorrhea prevalence. Within each uptake level, relative to the cumulative number of infections when DoxyPEP was implemented universally, targeting only the high-risk group increased cumulative infections by a percent change of 4.1-6.8% at 5 years and 0.3-1.3% at 20 years. These differences in the cumulative number of infections are negligible compared to the difference in DoxyPEP doses that would be required to cover 10% versus 100% of the MSM population for up to 20 years, suggesting that reducing gonorrhea incidence in the high-risk core group indirectly benefited the entire population by reducing transmission. Restricting DoxyPEP use to the high activity group also extended the time until doxycycline resistance reached the 87% prevalence threshold across all uptake levels (Table 1, Supplementary Table 2). These findings support recommending DoxyPEP only for those at high-risk for bacterial STIs.

The success of DoxyPEP, in terms of its ability to effectively reduce the burden of gonococcal infection over time, was insensitive to the probability of *de novo* doxycycline resistance parameter associated with DoxyPEP use (Supplementary Figures 6-7). One plausible explanation is that doxycycline resistance is already so prevalent in the population at baseline (26.8%) that small contributions to resistance upon DoxyPEP use are insignificant in comparison. In contrast, the success of DoxyPEP was highly dependent on the fitness cost associated with doxycycline resistance (Supplementary Figures 6-7). High fitness costs (≥15%) paired with sufficient DoxyPEP uptake (≥ 25%) led to sustained periods of dramatically reduced gonorrhea prevalence relative to baseline. In other words, high fitness costs for doxycycline resistant strains meant that DoxyPEP could more successfully reduce gonorrhea infections and do this for a longer time prior to resistance-related failure. However, as of 2018, the estimated proportion of gonococcal infections with maintained tetracycline resistance in the U.S. MSM population was substantial (26.8%) (13). This suggests resistance does not incur a high fitness cost, as tetracyclines are not included in national treatment guidelines and are therefore not directly exerting selective pressure.

Interpretation of long-term model projections warrants caution. We assume, within each DoxyPEP uptake level, that ceftriaxone and DoxyPEP use stay constant over time. Beyond the point at which doxycycline resistance reaches 87% prevalence, and DoxyPEP is therefore < 10% effective in preventing infection (assuming κ = 0.25), it is unlikely that DoxyPEP use would be maintained. Similarly, per WHO recommendations, once ceftriaxone resistance reaches 5% prevalence, treatment protocols require revision. Changes in treatment regimens would impact selective pressures and could alter the prevalence of gonorrhea that the model re-equilibrates to in the future following widespread DoxyPEP failure. Future projections also rely on current gonorrhea dynamics being maintained, when in fact new tools for gonorrhea management and prevention (e.g., rapid antimicrobial susceptibility diagnostics and vaccines) in development have the potential to disrupt dynamics within the next 20 years.

We assume that resistance mechanisms to ceftriaxone and doxycycline are independent. While non-U.S. data suggests ceftriaxone reduced susceptibility is associated with chromosomally-mediated tetracycline resistance, the rarity of detected ceftriaxone reduced susceptibility in the U.S. limits our ability to study this association (13). If ceftriaxone resistance is more likely to emerge in doxycycline-resistant strains, or if failed DoxyPEP followed by treatment with ceftriaxone for active infection fosters an environment conducive to dual resistance emergence, then the time to widespread DoxyPEP failure would be accelerated as dual resistance would emerge and spread more quickly.

Using a mathematical model to approximate the complex dynamics of sexual behavior and gonorrhea transmission forces limiting assumptions (8). The model is calibrated using MLE to a single mean equilibrium prevalence target (3.0%). This value does not reflect our uncertainty in the true prevalence of gonococcal infection in the U.S. MSM population, as the ability for asymptomatic cases to go undetected makes prevalence difficult to estimate.

Estimating multiple parameters using a single prevalence target also means that individual model parameters are non-identifiable or cannot be obtained with precision. Therefore, results should not be interpreted as absolute predictions, but as projections of relative gonococcal infection and resistance dynamics under a set of explicit model parameterizations. Model output is best interpreted relatively for comparing the effects of different levels of DoxyPEP uptake to the baseline scenario with 0% DoxyPEP uptake, consistent with our presentation of prevalence ratios, incidence rate ratios, and percent differences.

The model ignores the potential for bystander selection, that is, selection experiences by *N. gonorrhoeae* attributable to treatment with ceftriaxone or doxycycline for other, co-occurring indications. One study estimates that bystander experiences comprise 4.8% of *N. gonorrhoeae*’s ceftriaxone exposures and 25-29.7% of doxycycline exposures, but how substantially these bystander experiences contribute to resistance is not well understood (19,20). The model also excludes importation of drug resistant strains into the population. There is no heterogeneity in the modeled population except for sexual activity, defined by the annual rate of partner turnover, and sexual mixing assortativity by activity group. Infections are homogenous beyond symptom status and resistance profile; the model does not differentiate by anatomical site of infection.

DoxyPEP can achieve substantial reductions in gonorrhea prevalence and incidence in the short-term. However, the effectiveness of DoxyPEP for gonorrhea prophylaxis is limited by pre-existing *N. gonorrhoeae* doxycycline resistance, and its sustainability is limited by the selection for doxycycline resistant strains. Moreover, DoxyPEP does not appear to prolong the clinically useful lifespan of ceftriaxone monotherapy. Nonetheless, DoxyPEP’s clinical benefit could be deployed to temporarily minimize the burden of infection and disease as new tools are developed for gonorrhea prevention and treatment.

## Data Availability

All code needed to simulate the data, run analyses, and visualize results are available at https://github.com/emreichert13/doxypep.

## Ethical Approval

No ethical approval was required for this modeling study.

## Author Contributions

YHG can be credited with study conceptualization and supervision. ER developed the mathematical model, simulated modeled data, and conducted the analysis. ER wrote the original draft, and both authors participated in manuscript review and editing. ER and YHG had full access to and verified all data in the study. Both authors accept responsibility for the decision to submit for publication.

