## Supplementary Material for "Resistance and prevalence implications of doxycycline post-exposure prophylaxis for gonorrhea prevention in men who have sex with men: a modeling study"

**Supplementary Table 1.** Model parameters

| | Description | Value | Source(s) |
|---|---|---|---|
| *Model Calibration Target* | | | |
| | Gonorrhea prevalence at start (Calibration target mean) | 3.0% | |
| *Model Population, Sexual Behavior parameters* | | | |
| N | Population size | $10^6$ | Assumption |
| n | Relative size of sexual activity groups | | Tuite et al. 2017(15) |
| | Low | $n_1 = 0.3$ | |
| | Intermediate | $n_2 = 0.6$ | |
| | High | $n_3 = 0.1$ | |
| θ | Rate of partner turnover per sexual activity group (per year) | | Model fitting |
| | Low | $\theta_1 = 1*1.21$ | |
| | Intermediate | $\theta_2 = 5*1.21$ | |
| | High | $\theta_3 = 20*1.21$ | |
| ε | Mixing parameter | 0.24 | Model fitting |
| | Proportion of cases ceftriaxone resistant at start | 0.0001 | CDC GISP 2020-2021(14) |
| | Proportion of cases doxycycline resistant at start | 0.268 (0.20-0.80) | Mortimer and Grad 2023(13) (range) |
| ρ | Model entry/exit rate (per year) | 1/20 | Tuite et al. 2017 |
| *Gonorrhea Natural History parameters* | | | |
| σ | Proportion of **incident** infections that are symptomatic | 0.60 | Model fitting |
| b | Transmission probability per partnership | 0.46 | Model fitting |
| δ | Natural recovery rate from infection (per year) | 1/0.459 | Model fitting |

| | | | |
|---|---|---|---|
| $1/\gamma$ | Average time exposed, in days (independent of DoxyPEP treatment) | 1 | Assumption |
| *Treatment parameters* | | | |
| $T_s$ | Treatment rate, symptomatic infection (per year) | 1/0.031 | Model fitting |
| $T_{sr}$ | Retreatment rate, symptomatic infection (per year) | $T_S/3$ | Model fitting |
| $T_m$ | Screening rate, asymptomatic infection (per year) | 0.40 | Model fitting |
| $\xi_B$ | Proportion receiving DoxyPEP treatment upon exposure (i.e., uptake level) | 0-0.75 | Assumption, range explored in primary analysis |
| $\kappa$ | Proportion of DoxyPEP treatments that fail to prevent infection, for reasons not due to doxycycline resistance | 0.25 (0-0.8) | Assumption, Luetkemeyer et al. 2023(2) (range) |
| $\omega$ | Probability of emergence of resistance upon treatment | | |
| | Ceftriaxone | $\omega_A = 10^{-8}$ | Tuite et al. 2017, Vegvari et al. 2020(16) |
| | Doxycycline | $\omega_B = 0$ ($0\text{-}10^{-4}$) | Assumption (range) |
| f | Relative fitness of resistant bacteria, compared to susceptible | | |
| | Ceftriaxone resistant | $f_A = 0.98$ | Tuite et al. 2017 |
| | Doxycycline resistant | $f_B = 0.98$ (0.80-1) | Assumption (range) |
| | Dual resistance | $f_{AB} = f_A * f_B$ | Assumption |
| $\pi_s$ | Probability of retreatment if initial treatment failure, symptomatic infection | 0.90 | Tuite et al. 2017 |

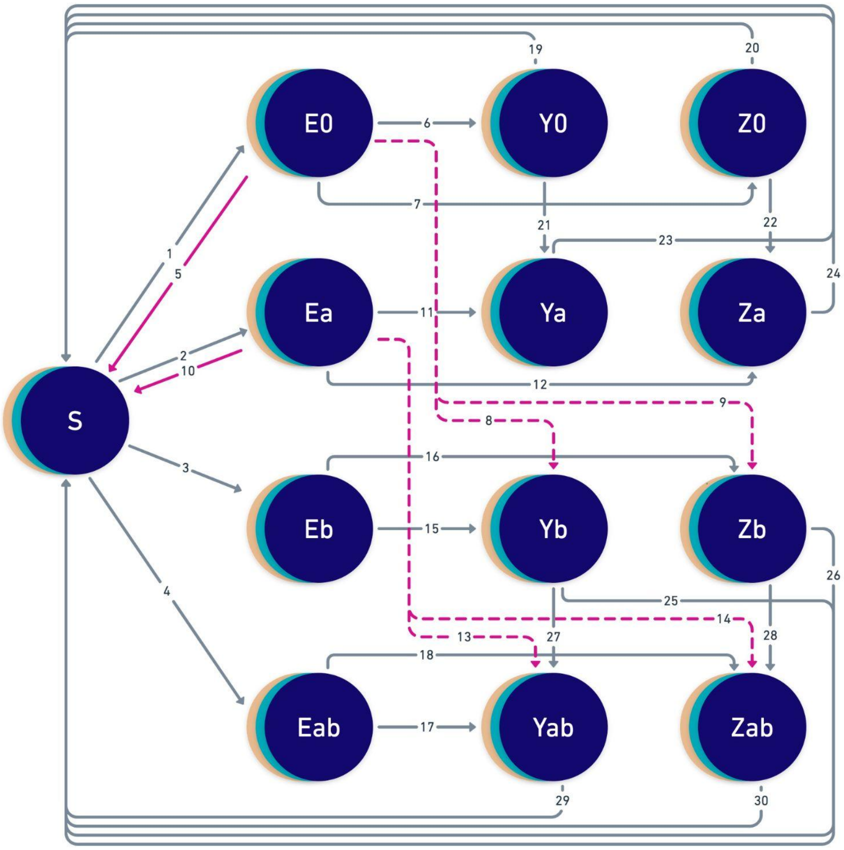


| | |
|---|---|
| 1 | $\boldsymbol{\beta}\,(\mathbf{Y}_0 + \mathbf{Z}_0)$ |
| 2 | $f_A \boldsymbol{\beta}\,(\mathbf{Y}_A + \mathbf{Z}_A)$ |
| 3 | $f_B \boldsymbol{\beta}\,(\mathbf{Y}_B + \mathbf{Z}_B)$ |
| 4 | $f_{AB} \boldsymbol{\beta}\,(\mathbf{Y}_{AB} + \mathbf{Z}_{AB})$ |
| 5, 10 | $\xi_B\,(1-\kappa)\,(1-\omega_B)\,\gamma$ |
| 6, 11 | $\sigma\gamma\,[(1-\xi_B) + \xi_B\,\kappa\,(1-\omega_B)]$ |
| 7, 12 | $(1-\sigma)\gamma\,[(1-\xi_B) + \xi_B\,\kappa\,(1-\omega_B)]$ |
| 8, 13 | $\sigma\xi_B\,\omega_B\,\gamma$ |
| 9, 14 | $(1-\sigma)\xi_B\,\omega_B\,\gamma$ |
| 15, 17 | $\sigma\gamma$ |
| 16, 18 | $(1-\sigma)\gamma$ |
| 19, 25 | $(1-\omega_A)\,T_s + \delta$ |
| 20, 26 | $(1-\omega_A)\,T_m + \delta$ |
| 21, 27 | $\omega_A T_s$ |
| 22, 28 | $\omega_A T_m$ |
| 23, 29 | $\pi_s T_{sr} + \delta$ |
| 24, 30 | $\delta$ |

**Supplementary Figure 1. Schematic of gonorrhea transmission model.** Abbreviations: S = Susceptible, E = Exposed, Y = Symptomatic Infection, Z = Asymptomatic Infection. Infections are further stratified by resistance profile, where 0 = susceptible, a = ceftriaxone resistant, b = doxycycline resistant, and ab = resistant to both drugs. Overlapping discs for all compartments represent the model's stratification into three sexual activity groups: low, intermediate, and high. Arrows depict rates between compartments and would be multiplied by the compartment from which they flow to generate the model's set of differential equations (see Technical Supplement). Arrows highlighted in pink represent transitions only possible with DoxyPEP implementation, and those that are dotted only occur if there is some non-zero probability of *de novo* resistance emerging with DoxyPEP use. Individuals can also enter and exit the population at rate ρ (arrows not shown). Definitions of all parameters used in rate equations can be found in Supplementary Table 1.

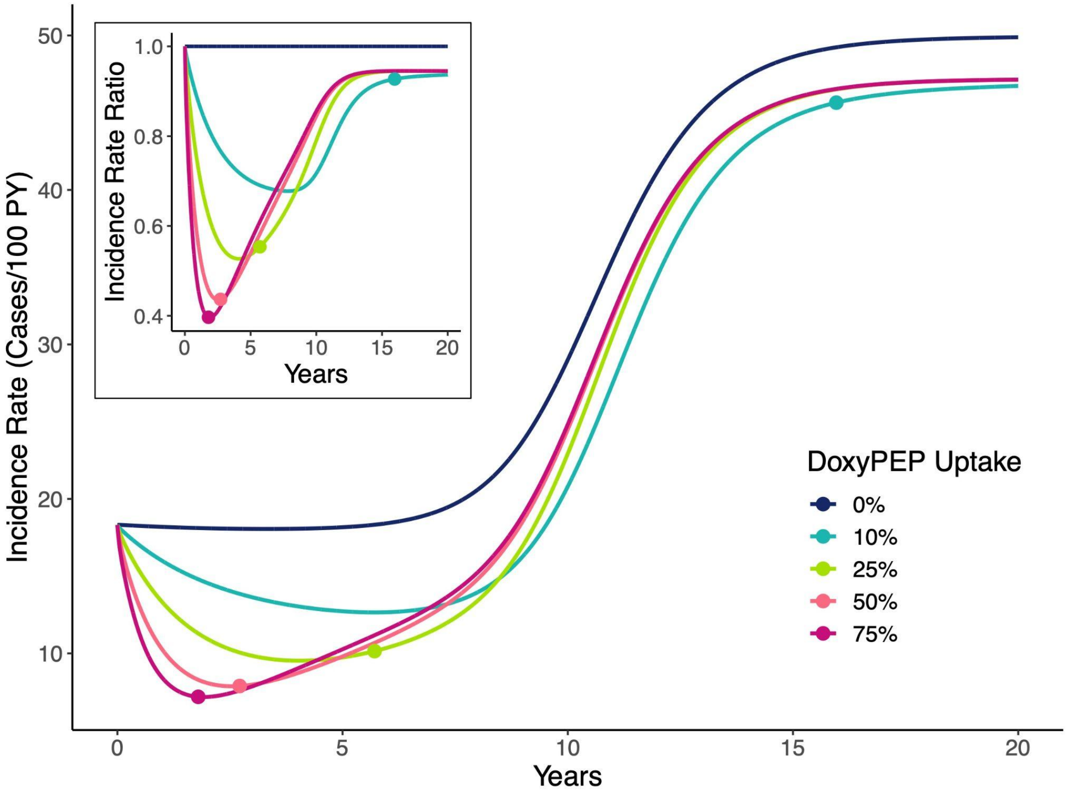


**Supplementary Figure 2. Incidence rate of gonococcal infection over time for varying DoxyPEP uptake levels (%).** The incident rate is calculated as the number of incident cases per 100 years of susceptible person-time. The upper left inset visualizes the same data as an incidence rate ratio (IRR), where results are normalized (or divided by) the incidence rate under the no DoxyPEP introduction scenario (0% uptake). Lines are colored by DoxyPEP uptake, defined as the proportion of exposure events that are treated with doxycycline post-exposure prophylaxis within the population. The points along these lines represent the time at which doxycycline resistance reached an 87% prevalence among gonococcal infections under that uptake level. Abbreviations: DoxyPEP = doxycycline post-exposure prophylaxis, PY = person-years at risk.

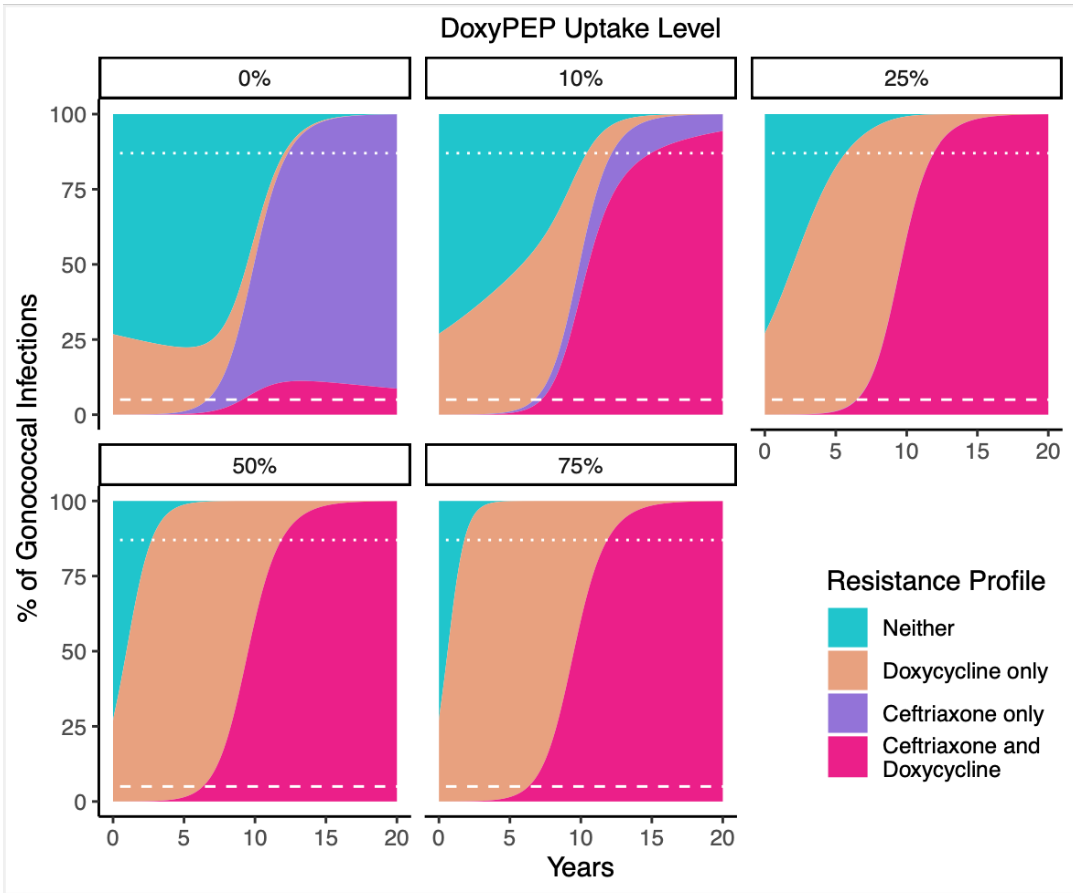


**Supplementary Figure 3. Percentage of gonococcal infections with each resistance profile over time, by DoxyPEP uptake (%).** Each facet depicts a different DoxyPEP uptake level. White dashed lines indicate the 5% threshold where WHO recommends changing treatment strategies (most relevant for ceftriaxone), and white dotted lines indicate the upper 87% threshold that may warrant discontinuation of DoxyPEP due to widespread doxycycline resistance.

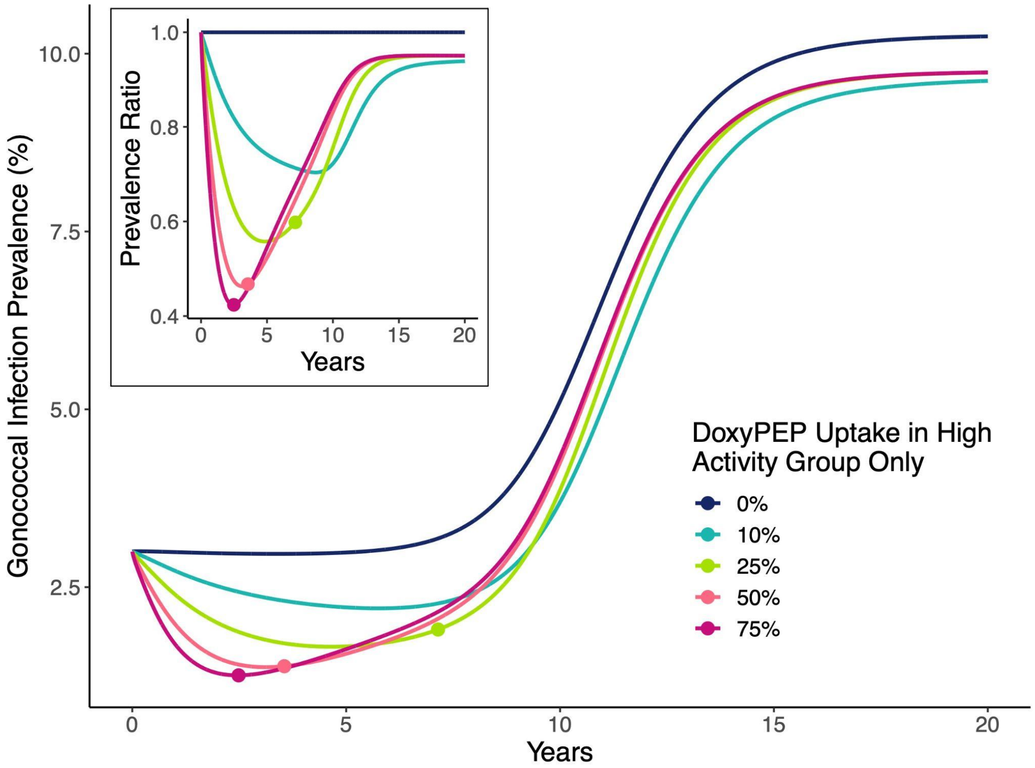


**Supplementary Figure 4. Prevalence of gonococcal infection over time for varying DoxyPEP uptake levels (%) in the high sexual activity group only.** Prevalence at each timepoint is calculated as the total number of infections over the total population size (N = $10^6$). The upper left inset visualizes the same data as a prevalence ratio (PR), where results are normalized (or divided by) the prevalence under the no DoxyPEP introduction scenario (0% uptake). Lines are colored by DoxyPEP uptake, defined as the proportion of exposure events that are treated with doxycycline post-exposure prophylaxis within the high activity group only. DoxyPEP uptake in the low and intermediate activity groups is fixed at 0%. The points along these lines represent the time at which doxycycline resistance met an 87% prevalence among gonococcal infections under that uptake level. Abbreviations: DoxyPEP = doxycycline post-exposure prophylaxis.

**Supplementary Table 2.** Clinically relevant endpoints by level of DoxyPEP uptake (%) in the high sexual activity group only, with DoxyPEP uptake in the low and intermediate activity groups fixed at 0%. Results are calculated at the total population level using baseline model parameters listed in Supplementary Table 1.

| DoxyPEP Uptake in High Activity Group Only | Time to Ceftriaxone Resistance at 5% (years) | Time to Dual Resistance at 5% (years) | Time to Doxycycline Resistance at 87% (years) | Reduction in Ceftriaxone Treatments at 5 years (%), relative to no DoxyPEP uptake | Reduction in Ceftriaxone Treatments at 20 years (%), relative to no DoxyPEP uptake | Reduction in Cumulative Infections at 5 years (%), relative to no DoxyPEP uptake | Reduction in Cumulative Infections at 20 years (%), relative to no DoxyPEP uptake |
|---|---|---|---|---|---|---|---|

| | | | | | | | |
|---|---|---|---|---|---|---|---|
| 0% | 6.6 | 9.1 | *NA* | *NA* | *NA* | *NA* | *NA* |
| 10% | 6.7 | 7.4 | *NA* | 17.8% | 13.9% | 16.2% | 12.3% |
| 25% | 6.5 | 6.6 | 7.1 | 34.5% | 16.4% | 32.2% | 14.1% |
| 50% | 6.3 | 6.3 | 3.6 | 46.1% | 16.8% | 43.5% | 14.3% |
| 75% | 6.3 | 6.3 | 2.5 | 50.0% | 16.9% | 47.5% | 14.3% |

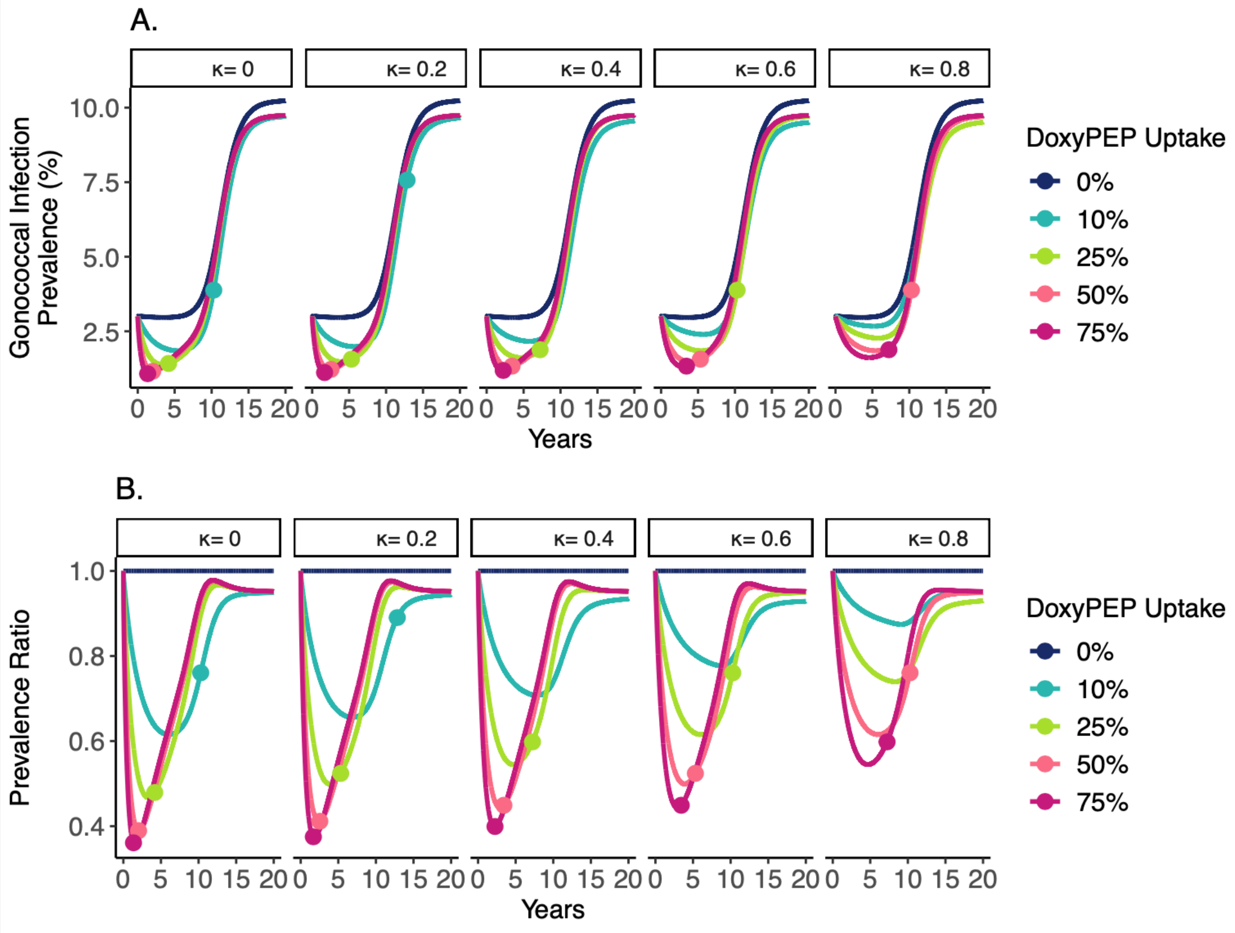


**Supplementary Figure 5. Prevalence of gonococcal infection over time for varying DoxyPEP uptake levels (%), by the per exposure risk ratio of infection for DoxyPEP users relative to non-users exposed to a doxycycline susceptible strain. A)** Absolute prevalence estimates over time, calculated as the total number of gonococcal infections over the total population size (N = $10^6$) under each DoxyPEP utilization scenario. **B)** Prevalence ratio (PR) estimates over time, where results are normalized (or divided by) the prevalence under the no DoxyPEP introduction scenario (0% uptake). Vertical facets (columns) show the risk ratio, also explained as the proportion of DoxyPEP treatments that fail to prevent infection for reasons not due to doxycycline resistance (κ). This is relative to the risk of infection for individuals exposed that are not using DoxyPEP (risk ratio = 1). Lines are colored by DoxyPEP uptake, defined as the proportion of exposure events that are treated with doxycycline post-exposure prophylaxis within the population. The points along these lines represent the time at which doxycycline resistance has reached an 87% prevalence among gonococcal infections under that uptake level. Abbreviations: DoxyPEP = doxycycline post-exposure prophylaxis.

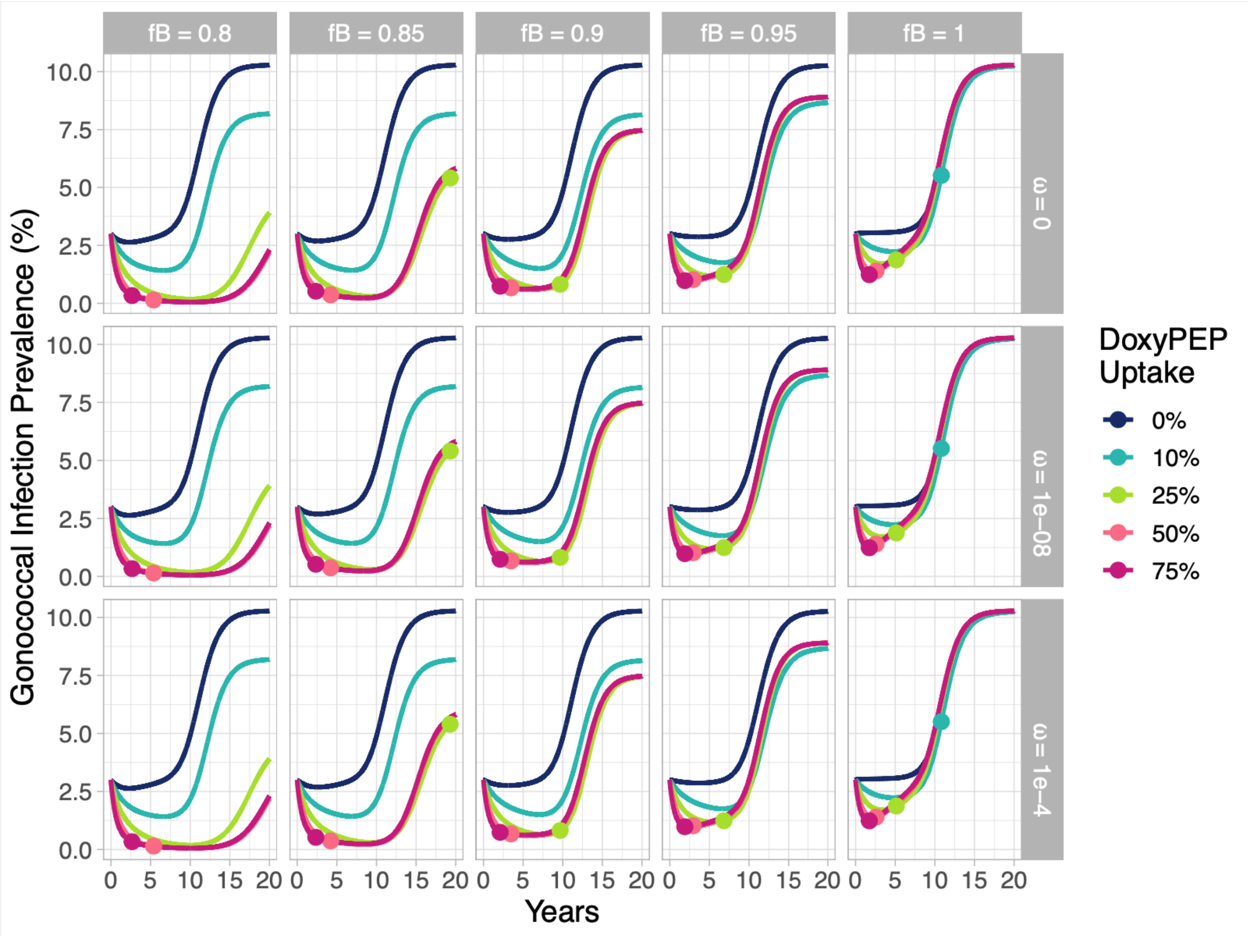


**Supplementary Figure 6. Prevalence of gonococcal infection over time for varying DoxyPEP uptake levels (%), by properties of doxycycline.** Vertical facets (columns) show the relative fitness of doxycycline resistant strains relative to susceptible strains ($f_B$), and horizontal facets (rows) represent probabilities of *de novo* resistance emergence upon DoxyPEP treatment ($\omega_B$). Lines are colored by DoxyPEP uptake, defined as the proportion of exposure events that are treated with doxycycline post-exposure prophylaxis within the population. The points along these lines represent the time at which doxycycline resistance has reached an 87% prevalence among gonococcal infections under that uptake level. Abbreviations: DoxyPEP = doxycycline post-exposure prophylaxis.

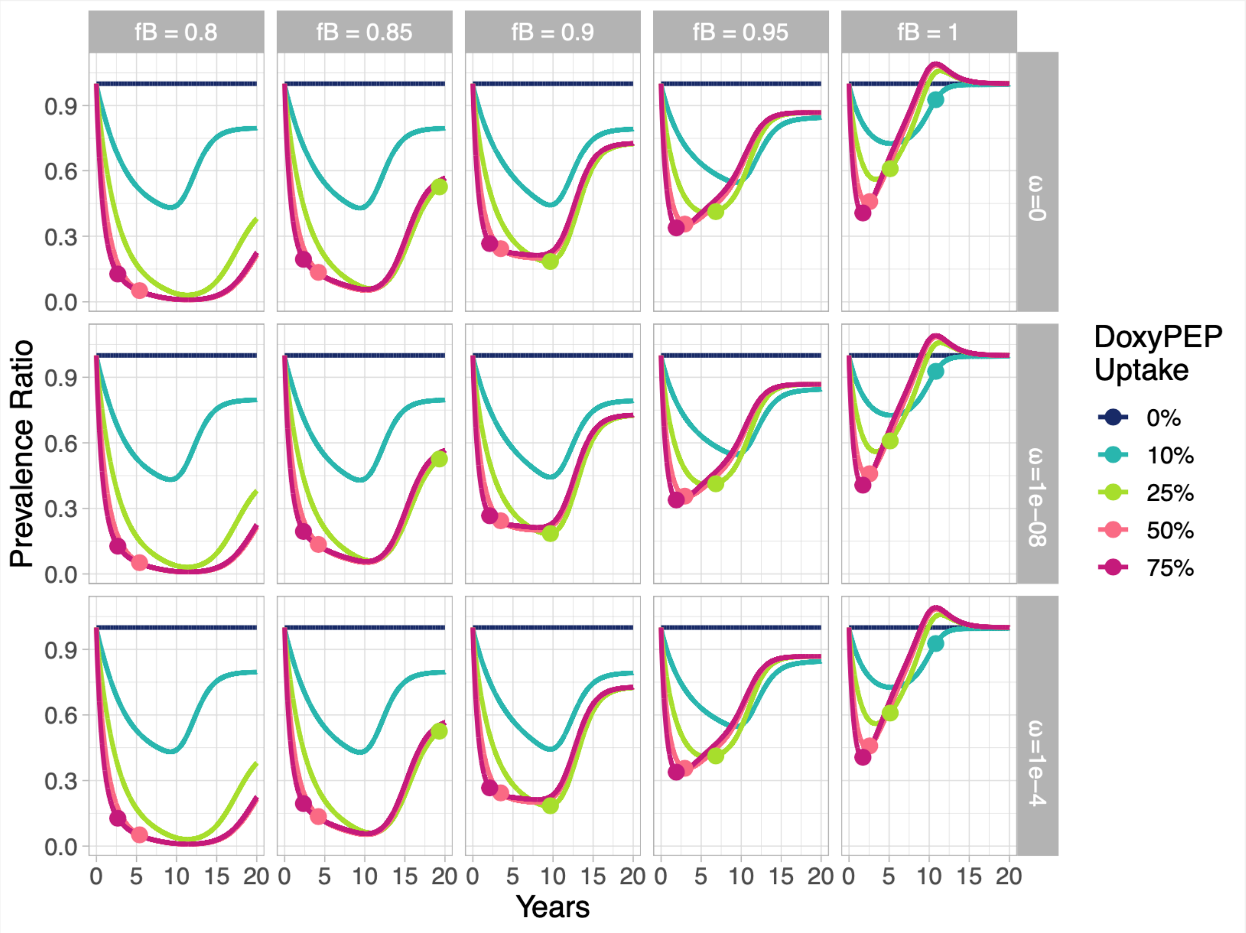


**Supplementary Figure 7. Prevalence ratio of gonococcal infection over time for varying DoxyPEP uptake levels (%) relative to no DoxyPEP uptake, by properties of doxycycline.** Vertical facets (columns) show the relative fitness of doxycycline resistant strains relative to susceptible strains ($f_B$), and horizontal facets (rows) represent probabilities of *de novo* resistance emergence upon DoxyPEP treatment ($\omega_B$). Lines are colored by DoxyPEP uptake, defined as the proportion of exposure events that are treated with doxycycline post-exposure prophylaxis within the population. The prevalence ratio is calculated at each timepoint by dividing the prevalence of gonococcal infection under each DoxyPEP uptake scenario by the prevalence given 0% DoxyPEP uptake. The point along each line represents the time at which doxycycline resistance has reached an 87% prevalence among gonococcal infections under that uptake level. Abbreviations: DoxyPEP = doxycycline post-exposure prophylaxis.

### Technical Supplement

### Model Structure

We adapted a single sex compartmental gonorrhea transmission model[1]. A visual overview of the susceptible-exposed-infectious-susceptible (SEIS) model is presented in Supplementary Figure 1. As previously described[1], infections are stratified by symptomatic ($Y$) versus asymptomatic ($Z$) infection, as well as by resistance profile, where each infection can be caused by bacteria resistant to ceftriaxone ($A$), doxycycline ($B$), neither, or both. The total size of the population ($N$) is set at $10^6$ and the absolute size of each sexual activity group at $N_k$, with the relative size of each group fixed at $n_k$. Therefore,

$$N_k = S_k + E_k + I_k, \text{ where}$$
$$E_k = E_{0k} + E_{Ak} + E_{Bk} + E_{ABk}, \text{ and}$$
$$I_k = Y_{0k} + Y_{Ak} + Y_{Bk} + Y_{ABk} + Z_{0k} + Z_{Ak} + Z_{Bk} + Z_{ABk}$$

The relative rate of partner change ($r_k$) for each sexual activity group is drawn from estimates by Tuite et al.[2] determined by data from the National HIV Behavioral Surveillance System[3]. The rate of partner change ($c_{min}$) in the low activity group was estimated through the maximum likelihood estimation (MLE) model fitting procedure. The annual rate of partner change for each activity group ($\theta_k$) is therefore described by the equation:

$$\theta_k = r_k * c_{min}$$

Assortativity between sexual activity groups is characterized by mixing parameter $\varepsilon$ [which can range between 0 (random mixing) and 1 (fully assortative mixing) and was determined through model fitting]. This leads the probability of an individual from sexual activity group $i$ coming into sexual contact with an individual from group $j$ to be:

$$p_{ij} = \varepsilon x_{ij}\frac{\theta_i}{N_i} + (1-\varepsilon)\frac{\theta_i\theta_j}{\sum_{k=1}^{3}\theta_k N_k}$$

where $x_{ij}$ is equal to 1 if $i = j$ and equal to 0 if $i \neq j$. The rate of infection for susceptible individuals in sexual activity group $i$ from infected partners of group $j$ ($\beta_{i \leftarrow j}$) is proportional to this per capita probability of sexual contact between groups $i$ and $j$ ($p_{ij}$), as well as the transmission probability per partnership ($b$). This relationship is defined by:

$$\beta_{i \leftarrow j} = bp_{ij}$$

Therefore, the per capita transmission matrix $\boldsymbol{\beta}$ is a $k \times k$ matrix with elements $\beta_{i \leftarrow j}$ at row i and column j, defined as the per capita rate of gonorrhea transmission to group i from group j. Note that the transmission matrix at element $\beta_{i \leftarrow j}$ is divided through by the size of group j's population, which stays constant over time. As area is also held constant, and deemed not influential in per capita transmission rates, the model assumes constant density and yields results identical to frequency-dependent transmission.

**Model Recovery from Gonococcal Infection**

Parameters describing treatment and retreatment rates by infection type (symptomatic vs. asymptomatic), as well as the rate of natural clearance of infection, are presented in Supplementary Table 1. These parameters can be used to calculate the average duration of an infection (years), which is ($1/T_s$) for treated symptomatic infections, ($1/T_m$) for treated asymptomatic infections detected via screening, and ($1/\delta$) for infections that clear naturally. The treatment rate $T_{sr}$ for those with an initial treatment failure assumes an average duration of infection three times that of those with initial treatment success; $1/T_{sr}$ represents the average time in years until successful retreatment, including the time it takes for: 1) the individual to receive the initial failed treatment, 2) the individual to re-seek care, and 3) the provider to identify and prescribe the correct antibiotic for retreatment. It is possible for resistance acquired upon initial treatment of a susceptible infection to then be retreated insufficiently with the same antibiotic before receiving a successful retreatment; in these instances, the average duration of infection is ($1/T_s + 1/T_{sr}$), in years.

**Model Equations**

In matrix form, the model is described by the following system of differential equations, where ◦ denotes element-wise multiplication. All parameters are defined in Supplementary Table 1. Bolded letters represent $k \times 1$ column vectors with compartmental variables for each sexual activity group (e.g., $\mathbf{S} = [S_1,...S_k]^T$) for $k$ groups), with the exception of the $k \times k$ matrix $\boldsymbol{\beta}$.

$$d\mathbf{S}/dt = -\boldsymbol{\beta}((\mathbf{Y}_0 + \mathbf{Z}_0) + f_A(\mathbf{Y}_A + \mathbf{Z}_A) + f_B(\mathbf{Y}_B + \mathbf{Z}_B) + f_{AB}(\mathbf{Y}_{AB} + \mathbf{Z}_{AB}))\circ\mathbf{S} +$$
$$(1-\omega_A)(T_s\mathbf{Y}_0 + T_m\mathbf{Z}_0) +$$
$$\pi_s T_{sr}(\mathbf{Y}_A + \mathbf{Y}_{AB}) +$$
$$(1-\omega_A)(T_s\mathbf{Y}_B + T_m\mathbf{Z}_B) +$$
$$\xi_B(1-\kappa)(1-\omega_B)\gamma(\mathbf{E}_0 + \mathbf{E}_A) +$$
$$\delta(\mathbf{Y}_0 + \mathbf{Y}_A + \mathbf{Y}_B + \mathbf{Y}_{AB} + \mathbf{Z}_0 + \mathbf{Z}_A + \mathbf{Z}_B + \mathbf{Z}_{AB}) +$$
$$\rho\mathbf{N} - \rho\mathbf{S}$$

$$d\mathbf{E}_0/dt = \boldsymbol{\beta}(\mathbf{Y}_0 + \mathbf{Z}_0)\circ\mathbf{S} - \gamma\mathbf{E}_0$$
$$d\mathbf{Y}_0/dt = \sigma(1-\xi_B)\gamma\mathbf{E}_0 + \sigma\xi_B\kappa(1-\omega_B)\gamma\mathbf{E}_0 - T_s\mathbf{Y}_0 - \delta\mathbf{Y}_0 - \rho\mathbf{Y}_0$$
$$d\mathbf{Z}_0/dt = (1-\sigma)(1-\xi_B)\gamma\mathbf{E}_0 + (1-\sigma)\xi_B\kappa(1-\omega_B)\gamma\mathbf{E}_0 - T_m\mathbf{Z}_0 - \delta\mathbf{Z}_0 - \rho\mathbf{Z}_0$$

$$d\mathbf{E}_A/dt = f_A\boldsymbol{\beta}(\mathbf{Y}_A + \mathbf{Z}_A)\circ\mathbf{S} - \gamma\mathbf{E}_A$$
$$d\mathbf{Y}_A/dt = \sigma(1-\xi_B)\gamma\mathbf{E}_A + \sigma\xi_B\kappa(1-\omega_B)\gamma\mathbf{E}_A + \omega_A T_s\mathbf{Y}_0 + \omega_A T_s\mathbf{Y}_0 - \pi_s T_{sr}\mathbf{Y}_A - \delta\mathbf{Y}_A - \rho\mathbf{Y}_A$$
$$d\mathbf{Z}_A/dt = (1-\sigma)(1-\xi_B)\gamma\mathbf{E}_A + (1-\sigma)\xi_B\kappa(1-\omega_B)\gamma\mathbf{E}_A + \omega_A T_m\mathbf{Z}_0 - \delta\mathbf{Z}_A - \rho\mathbf{Z}_A$$

$$d\mathbf{E}_B/dt = f_B\boldsymbol{\beta}(\mathbf{Y}_B + \mathbf{Z}_B)\circ\mathbf{S} - \gamma\mathbf{E}_B$$

$d\boldsymbol{Y}_B/dt = \sigma\gamma\boldsymbol{E}_B + \sigma\xi_B\omega_B\gamma\boldsymbol{E}_0 - T_s\boldsymbol{Y}_B - \delta\boldsymbol{Y}_B - \rho\boldsymbol{Y}_B$
$d\boldsymbol{Z}_B/dt = (1\text{-}\sigma)\gamma\boldsymbol{E}_B + (1\text{-}\sigma)\xi_B\omega_B\gamma\boldsymbol{E}_0 - T_m\boldsymbol{Z}_B - \delta\boldsymbol{Z}_B - \rho\boldsymbol{Z}_B$

$d\boldsymbol{E}_{AB}/dt = f_{AB}\boldsymbol{\beta}(\boldsymbol{Y}_{AB} + \boldsymbol{Z}_{AB})\circ\boldsymbol{S} - \gamma\boldsymbol{E}_{AB}$
$d\boldsymbol{Y}_{AB}/dt = \sigma\gamma\boldsymbol{E}_{AB} + \sigma\xi_B\omega_B\gamma\boldsymbol{E}_A + \omega_A T_s\boldsymbol{Y}_B - \pi_s T_{sr}\boldsymbol{Y}_{AB} - \delta\boldsymbol{Y}_{AB} - \rho\boldsymbol{Y}_{AB}$
$d\boldsymbol{Z}_{AB}/dt = (1\text{-}\sigma)\gamma\boldsymbol{E}_{AB} + (1\text{-}\sigma)\xi_B\omega_B\gamma\boldsymbol{E}_A + \omega_A T_m\boldsymbol{Z}_B - \delta\boldsymbol{Z}_{AB} - \rho\boldsymbol{Z}_{AB}$

We calculate the number of incident infections at time *t*, or the overall force of infection across sexual activity groups, as $\lambda_t = \boldsymbol{\beta}((\boldsymbol{Y}_{0t} + \boldsymbol{Z}_{0t}) + f_A(\boldsymbol{Y}_{At} + \boldsymbol{Z}_{At}) + f_B(\boldsymbol{Y}_{Bt} + \boldsymbol{Z}_{Bt}) + f_{AB}(\boldsymbol{Y}_{ABt} + \boldsymbol{Z}_{ABt}))\circ\boldsymbol{S}_t$.

The overall prevalence of infection at time t can be calculated as $Prev_t = (\boldsymbol{Y}_{0t} + \boldsymbol{Z}_{0t} + \boldsymbol{Y}_{At} + \boldsymbol{Z}_{At} + \boldsymbol{Y}_{Bt} + \boldsymbol{Z}_{Bt} + \boldsymbol{Y}_{ABt} + \boldsymbol{Z}_{ABt})/N$.


1. Reichert E, Yaesoubi R, Rönn MM, Gift TL, Salomon JA, Grad YH. Resistance-minimizing strategies for introducing a novel antibiotic for gonorrhea treatment: a mathematical modeling study. medRxiv [Internet]. medRxiv: medRxiv; 2023 [cited 2023 Mar 27]. p. 2023.02.14.23285710. Available from: https://www.medrxiv.org/content/10.1101/2023.02.14.23285710v1
2. Tuite AR, Gift TL, Chesson HW, Hsu K, Salomon JA, Grad YH. Impact of Rapid Susceptibility Testing and Antibiotic Selection Strategy on the Emergence and Spread of Antibiotic Resistance in Gonorrhea. *J Infect Dis*. 2017;216(9):1141-1149. doi:10.1093/infdis/jix450
3. Centers for Disease Control and Prevention. HIV Infection Risk, Prevention, and Testing Behaviors among Men Who Have Sex With Men -- National HIV Behavioral Surveillance, 20 U.S. Cities, 2014. HIV Surveillance Special Report 15. Published January 2016. Accessed September 8, 2022. http://www.cdc.gov/hiv/library/reports/surveillance/#panel2
